# COVID-19 Pandemic Prediction for Hungary; a Hybrid Machine Learning Approach

**DOI:** 10.1101/2020.05.02.20088427

**Authors:** Gergo Pinter, Imre Felde, Amir Mosavi, Pedram Ghamisi, Richard Gloaguen

## Abstract

Several epidemiological models are being used around the world to project the number of infected individuals and the mortality rates of the COVID-19 outbreak. Advancing accurate prediction models is of utmost importance to take proper actions. Due to a high level of uncertainty or even lack of essential data, the standard epidemiological models have been challenged regarding the delivery of higher accuracy for long-term prediction. As an alternative to the susceptible-infected-resistant (SIR)-based models, this study proposes a hybrid machine learning approach to predict the COVID-19 and we exemplify its potential using data from Hungary. The hybrid machine learning methods of adaptive network-based fuzzy inference system (ANFIS) and multi-layered perceptron-imperialist competitive algorithm (MLP-ICA) are used to predict time series of infected individuals and mortality rate. The models predict that by late May, the outbreak and the total morality will drop substantially. The validation is performed for nine days with promising results, which confirms the model accuracy. It is expected that the model maintains its accuracy as long as no significant interruption occurs. Based on the results reported here, and due to the complex nature of the COVID-19 outbreak and variation in its behavior from nation-to-nation, this study suggests machine learning as an effective tool to model the outbreak. This paper provides an initial benchmarking to demonstrate the potential of machine learning for future research.

## 1 Introduction

Severe acute respiratory syndrome coronavirus 2, also known as SARS-CoV-2, is reported as a virus strain causing the respiratory disease of COVID-19 [1]. The World Health Organization (WHO) and the global nations confirmed the coronavirus disease to be extremely contagious [2,3]. The COVID-19 pandemic has been widely recognized as a public health emergency of international concern [4]. To estimate the outbreak, identify the peak ahead of time, and also predict the mortality rate the epidemiological models had been widely used by officials and media. Outbreak prediction models have shown to be essential to communicate insights into the likely spread and consequences of COVID-19. Furthermore, governments and other legislative bodies used the insights from prediction models to suggest new policies and to assess the effectiveness of the enforced policies [5].

The COVID-19 pandemic has been reported to be extremely aggressive to spread [6]. Due to the complex nature of the COVID-19 outbreak and its irregularity in different countries, the standard epidemiological models, i.e., susceptible-infected-resistant (SIR)-based models, had been challenged for delivering higher performance in individual nations. Furthermore, as the COVID-19 outbreak showed significant differences with other recent outbreaks, e.g., Ebola, Cholera, swine fever, H1N1 influenza, dengue fever, and Zika, advanced epidemiological models have been emerged to provide higher accuracy [7]. Nevertheless, due to several unknown variables involved in the spread, the complexity of population-wide behavior in various countries, and differences in containment strategies model uncertainty has been reported inevitable [8–10]. Consequently, standard epidemiological models face new challenges to deliver more reliable results.

The strategy standard SIR models is formed around the assumption of transmitting the infectious disease through contacts, considering three different classes susceptible, infected, and recovered [11]. The susceptible to infection (class S), infected (class I), and the removed population (class R) build the foundation of the epidemiological modeling. Note that definition of various classes of outbreak may vary. For instance, *R* is often referred to those that have recovered, developed immunity, been isolated, or passed away. However, in some countries, *R* is susceptible to be infected again and there exists uncertainties in allocating *R* a value. Advancing SIR-based models requires several assumptions. It is assumed that the class *I* transmits the infection to class *S* where the number of probable transmissions is proportional to the total number of contacts computed using basic differential equations as follows [12–14].

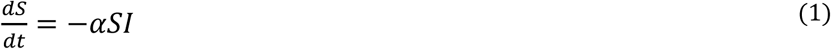

where *I*, *S*, and *α* represent the infected population, the susceptible population, and the daily reproduction rate, respectively. The value of *S* in the time-series produced by the differential equation gradually declines. At the early stage of the outbreak, it is assumed that *S* ≈ 1 where 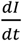 becomes linear. Eventually the class *I* can be stated as follows.

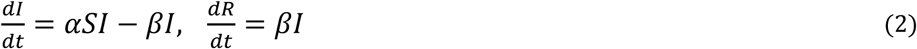

where *β* regulates the daily rate of spread. Furthermore, the individuals excluded from the model is computed as follows. Considering the above assumption, the outbreak modeling with SIR is finally computed as follows:

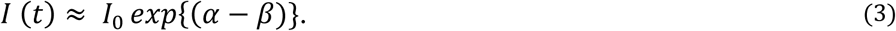

Furthermore, to evaluate the performance of the SIR-based models, the median success of the outbreak prediction is used which is calculated as follows:

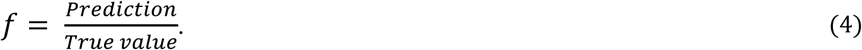

Several analytical solutions to the SIR models have been provided in literature [15,16]. As the different nations take different actions toward slowing down the outbreak, the SIR-based model must be adopted according to the local assumptions [17]. Inaccuracy of many SIR-based models in predicting the outbreak and mortality rate have been evidenced during the COVID-19 in many nations. The key success of an SIR-based model relies on choosing the right model according to the context and the relevant assumptions. SIS, SIRD, MSIR, SEIR, SEIS, MSEIR, and MSEIRS models are among the popular models used for predicting COVID-19 outbreaks worldwide. The more advanced variation of SIR-d models carefully considers the vital dynamics and constant population [16]. For instance, at the presence of the long-lasting immunity assumption when the immunity is not realized upon recovery from infection, the susceptible-infectious-susceptible (SIS) model was suggested [18]. In contrast, the susceptible-infected-recovered-deceased-model (SIRD) is used when immunity is assumed [19]. In the case of COVID-19 different nations took different approaches in this regard.

SEIR models have been reported among the most popular tools to predict the outbreak. SEIR models through considering the significant incubation period of an infected person reported to present relatively more accurate predictions. In the case of Varicella and Zika outbreaks the SEIR models showed increased model accuracy [20,21]. SEIR models assume that the incubation period is a random variable and similarly to the SIR model, there is a disease-free-equilibrium [22,23]. It should be noted, however, that SEIR models can not fit well where the contact network are non-stationary through time [24]. Social mixing as key factor of non-stationarity determines the reproductive number *R*_0_ which is the number of susceptible individuals for infection. The value of *R*_0_ for COVID-19 was estimated to be 4 which greatly trigged the pandemic [1]. The lockdown measures aimed at reducing the *R*_0_ value down to 1. Nevertheless, the SEIR models are reported to be difficult to fit in the case of COVID-19 due to the non-stationarity of mixing, caused by nudging intervention measures. Therefore, to develop accurate SIR-based models an in-depth information about the social movement and the quality of lockdown measures would be essential. Other drawback of SIR-based models is the short lead-time. As the lead-time increases, the accuracy of the model declines. For instance, for the COVID-19 outbreak in Italy, the accuracy of the model reduces from *f = 1* for the first five days to *f =* 0.86 for day 6 [17]. Overall, the SIR-based models would be accurate if firstly the status of social interactions is stable. Secondly, the class *R* can be computed precisely. To better estimate the class *R*, several data sources can be integrated with SIR-based models, e.g., social media and call data records (CDR), which of course a high degree of uncertainty and complexity still remains [25–32]. Considering the above uncertainties involved in the advancement of SIR-based models the generalization ability are yet to be improved to achieve scalable model with high performance [33].

Due to the complexity and the large-scale nature of the problem in developing epidemiological models, machine learning has recently gained attention for building outbreak prediction models. ML has already shown promising results in contribution to advancing higher generalization ability and greater prediction reliability for longer lead-times [34–38]. Machine learning has been already recognized a computing technique with great potential in outbreak prediction. Application of ML in outbreak prediction includes several algorithms, e.g., random forest for swine fever [39] [40], neural network for H1N1 flu, dengue fever, and Oyster norovirus [41] [11] [42], genetic programming for Oyster norovirus [43], classification and regression tree (CART) for Dengue [44], Bayesian Network for Dengue and Aedes [45], LogitBoost for Dengue [46], multi-regression and Naïve Bayes for Dengue outbreak prediction [47]. Although ML methods were used in modeling former pandemics (e.g., Ebola, Cholera, swine fever, H1N1 influenza, dengue fever, Zika, oyster norovirus [11,39–48]), there is a gap in the literature for peer-reviewed papers dedicated to COVID-19. Nevertheless, machine learning has been strongly proposed as a great potential for the fight against COVID-19 [49,50]. Machine learning delivered promising results in several aspects for mitigation and prevention and have been endorsed in the scientific community for, e.g., case identifications [51], classification of novel pathogens [52], modification of SIR-based models [53], diagnosis [54,55], survival prediction [56], and ICU demand prediction [57]. Furthermore, the non-peer reviewed sources suggest numerous potentials of machine learning to fight COVID-19. Among the applications of machine learning improvement of the existing models of prediction, identifying vulnerable groups, early diagnose, advancement of drugs delivery, evaluation of the probability of next pandemic, advancement of the integrated systems for spatio-temporal prediction, evaluating the risk of infection, advancing reliable biomedical knowledge graphs, and data mining the social networks are being noted.

As stated in our former paper machine learning can be used for data preprocessing. Improving the quality of data can particularly improve the quality of the SIR-based model. For instance, the number of cases reported by Worldometer is not precisely the number of infected cases (*E* in the SEIR model), or calculating the number of infectious people *(I* in SEIR) cannot be easily determined, as many people who might be infectious may not turn up for testing. Although the number of people who are admitted to hospital and deceased wont support *R* as most COVID-19 positive cases recover without entering hospital. Considering this data problem, it is extremely difficult to fit SEIR models satisfactorily. Considering such challenges, for future research, the ability of machine learning for estimation of the missing information on the number of exposed *E* or infecteds can be evaluated. Along with the prediction of the outbreak, prediction of the total mortality rate (*n*(deaths) / *n* (infecteds)) is also essential to accurately estimate the number of potential patients with the in the critical situation and the required beds in intensive care units. Although the research is in the very early stage, the trend in outbreak prediction with machine learning can be classified in two directions. Firstly, improvement of the SIR-based models, e.g., [53,58], and secondly time-series prediction [59,60]. Consequently, the state-of-the-art machine learning methods for outbreak modeling suggest two major research gaps for machine learning to address. Firstly, improvement of SIR-based models and secondly advancement in outbreak time series. Considering the drawbacks of the SIR-based models, machine learning should be able to contribute. This paper contributes to the advancement of time-series modelling and prediction of COVID-19. Although ML has long been established as a standard tool for modeling natural disasters and weather forecasting [61–65], its application in modeling outbreak is still in the early stages. More sophisticated ML methods are yet to be explored. A recent paper by Ardabili et al, [50], explored the potential of MLP and ANFIS in time series prediction of COVID-19 in several countries. Contribution of the present paper is to improve the quality of prediction through proposing a hybrid machine learning and compare the results with ANFIS. In the present paper the time series of the total mortality is also included.

The rest of this paper is organized as follows. Section two describes the methods and materials. The results are given in section three. Sections four presents conclusions.

## 2. Materials and methods

### 2.1 Data

Dataset is related to the statistical reports of COVID-19 cases and mortality rate of Hungary which is available at: https://www.worldometers.info/coronavirus/country/hungary/. Figure 1 and 2 presents the total and daily reports of COVID-19 statistics, respectively from 4-March to 19-April.

**Figure 1.**
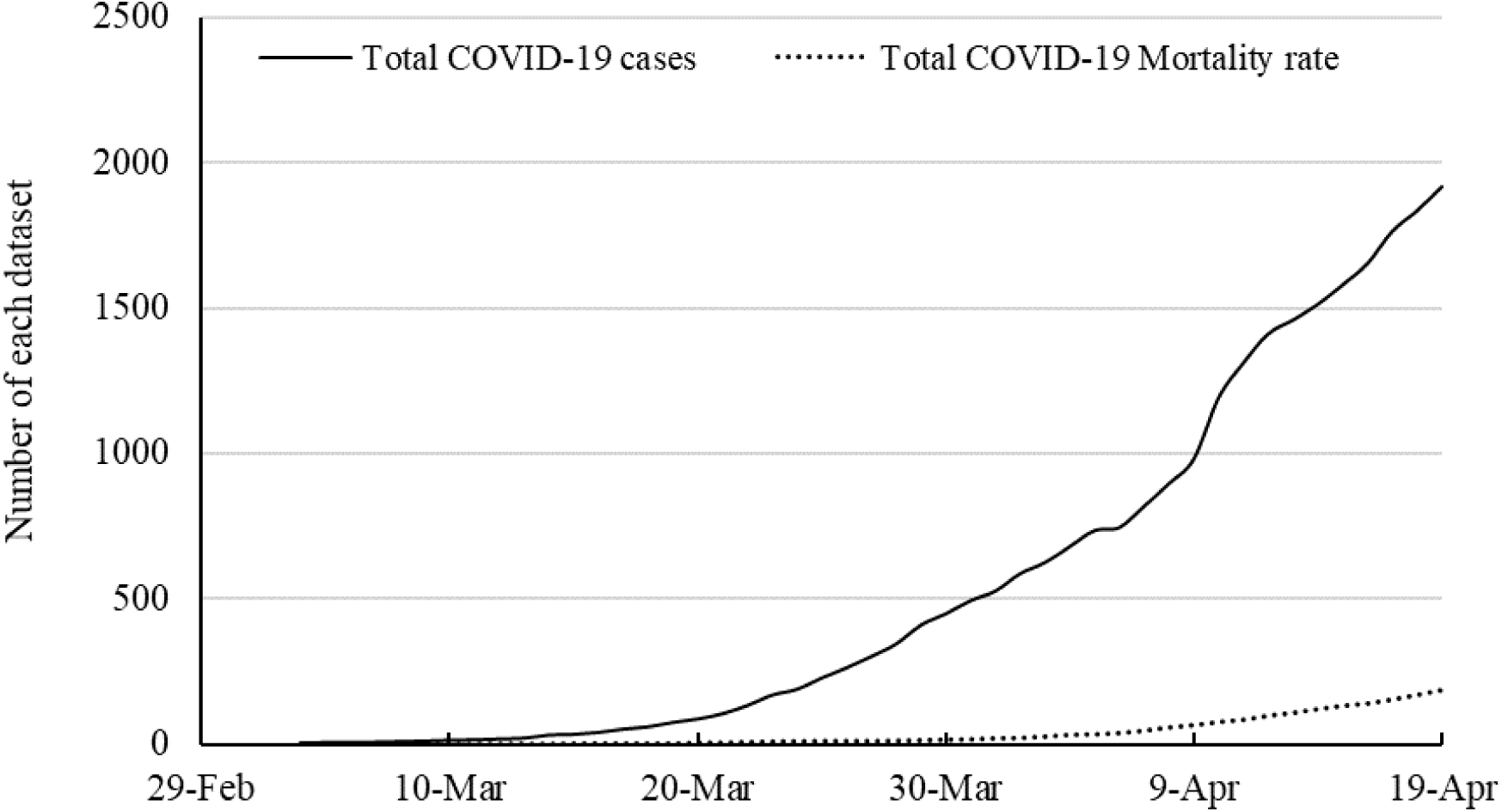
Total statistics for the number of cases and mortality rate of COVID-19

**Figure 2.**
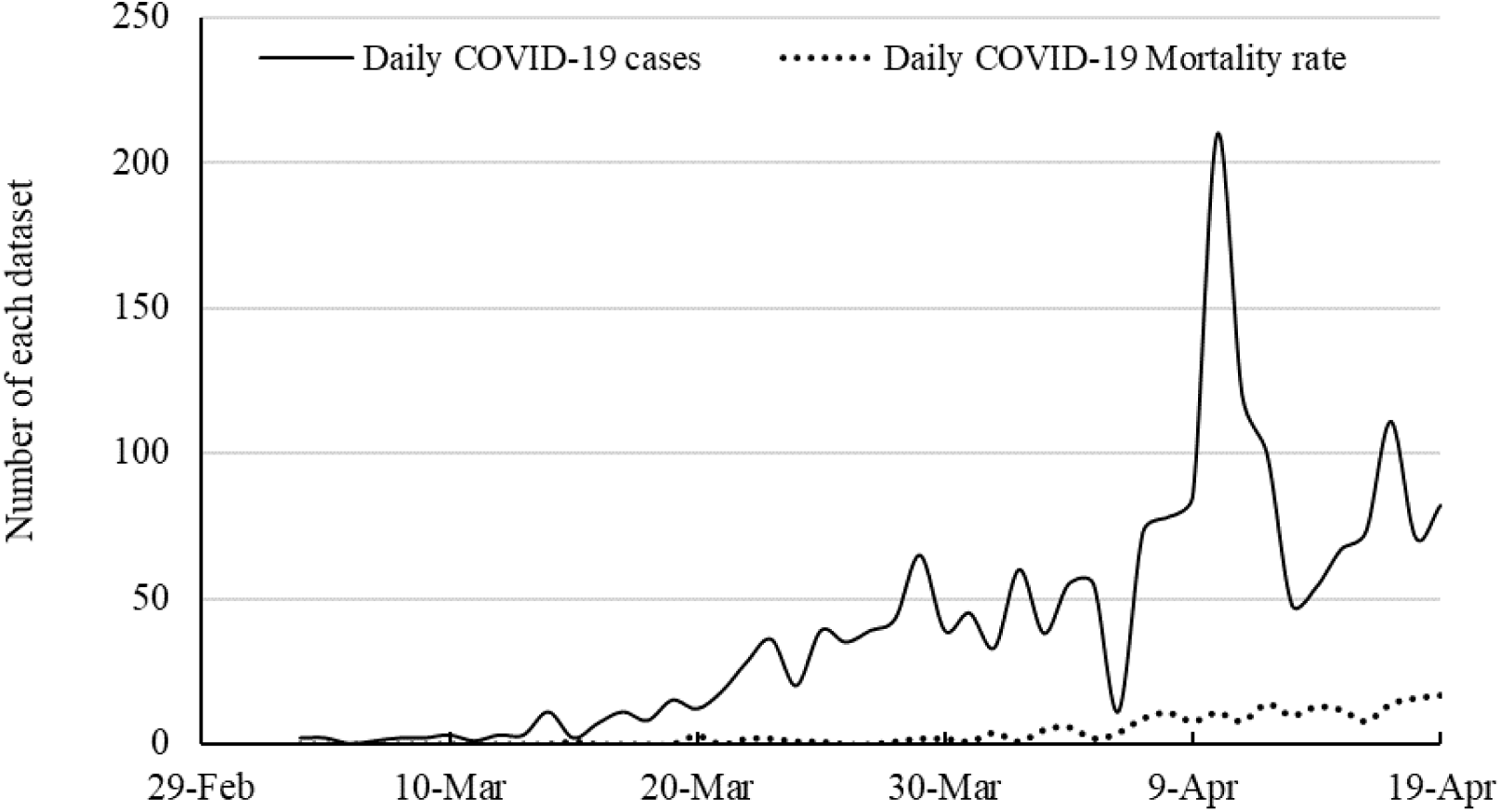
Daily statistics for the number of cases and mortality rate of COVID-19

### 2.2 Methods and modeling strategy

In the present study, modeling is performed by machine learning methods. Training is the basis of these methods as well as many artificial intelligence (AI) methods [66,67]. According to some psychologists, humans or living things interact with their surroundings by trial and error and achieve the best performance to reach a goal. Based on this theory and using the ability of computers to repeat a set of instructions, these conditions can be provided for computer programs to interact with the environment by updating values and optimizing functions, according to the results of interaction with the environment to solve a problem or achieve a specific goal. How to update values and parameters in successive repetitions by a computer is called a training algorithm [68–70]. One of these methods is Neural Networks (NN), according to which the modeling of the connection of neurons in the human brain, software programs were designed to solve various problems. To solve these problems, operational NN such as classification, clustering, or function approximation are performed using appropriate learning methods [71,72]. The training of the algorithm is the initial and the important step for developing a model [73,74]. Developing a predictive AI model requires a dataset categorized into two sections i.e. input(s) (as independent variable(s)) and output(s) (as dependent variable(s)) [75].

In the present study, time-series data have been considered as the independent variables for the prediction of COVID-19 cases and mortality rate (as dependent variables). Time-series dataset was prepared based on two scenarios as described in table 1. The first scenario categorizes the time-series data into four inputs for the last four consequently odd days’ cases or mortality rate for the prediction of x_t_ as the next day’s case or mortality rate, and the second scenario categorizes the time-series data into four inputs for the last four consequently even days’ cases or mortality rate for the prediction of x_t_ as the next day’s case or mortality rate.

**Table 1.** Two proposed scenarios for time-series prediction of COVID-19 in Hungary

|  | Inputs | Outputs |
| --- | --- | --- |
| Scenario 1 | $X_{t-1}, X_{t-3}, X_{t-5}$ and $X_{t-7}$ | $X_t$ |
| Scenario 2 | $X_{t-2}, X_{t-4}, X_{t-6}$ and $X_{t-8}$ | $X_t$ |

In the present study the two robust hybrid methods of ANN algorithm i.e. MLP-ICA and ANFIS have been employed for developing the required models. The dataset is devised into two parts. One part is devoted to training and the second part, i.e., 20-28 April, is devoted to model validation.

#### 2.2.1 Hybrid multi layered perceptron-imperialist competitive algorithm (MLP-ICA)

Multi-layered-perceptron (MLP) is the frequently used ANN method for the prediction and modeling purposes. This technique as a single method provides an acceptable accuracy for prediction tasks in simple and semi-compleX dataset. But, in case of doing modeling tasks in compleX dataset, there is a need for more robust techniques [76,77]. For this reason, hybrid methods have been growing up [76,77]. Hybrid methods contain a predictor and one or more optimizer [76,77]. The present study develops a hybrid MLP-ICA method as a robust hybrid algorithm for developing a platform for predicting the COVID-19 cases and mortality rate in Hungary. The ICA is a method in the field of evolutionary calculations that seeks to find the optimal answer to various optimization problems. This algorithm, by mathematical modeling, provides a socio-political evolutionary algorithm for solving mathematical optimization problems [78]. Like the all algorithms in this category, the ICA constitutes a primary set of possible answers. These answers are known as countries in the ICA. The ICA gradually improves the initial responses (countries) and ultimately provides the appropriate answer to the optimization problem [78,79].

The algorithms are based on the policy of assimilation, imperialist competition and revolution. This algorithm, by imitating the process of social, economic, and political development of countries and by mathematical modeling of parts of this process, provides operators in the form of a regular algorithm that can help to solve complex optimization problems [78,80]. In fact, this algorithm looks at the answers to the optimization problem in the form of countries and tries to gradually improve these answers during a repetitive process and eventually reach the optimal answer to the problem [78,80]. In the *Nvar* dimension optimization problem, a country is an array of *Nvar* × *1* length. This array is defined as follows:

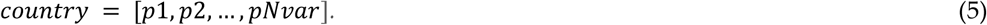

To start the algorithm, the country number of the initial country is created *(N_country_)*. To select the *N_imp_* as the best members of the population (countries with the lowest amount of cost function) as imperialists. The rest of the *N_col_* countries, form colonies belonging to an empire. To divide the early colonies between the imperialists, imperialist owns number of colonies, the number of which is proportional to its power [78,80]. The following figure symbolically shows how the colonies are divided among the colonial powers.

**Figure 3.**
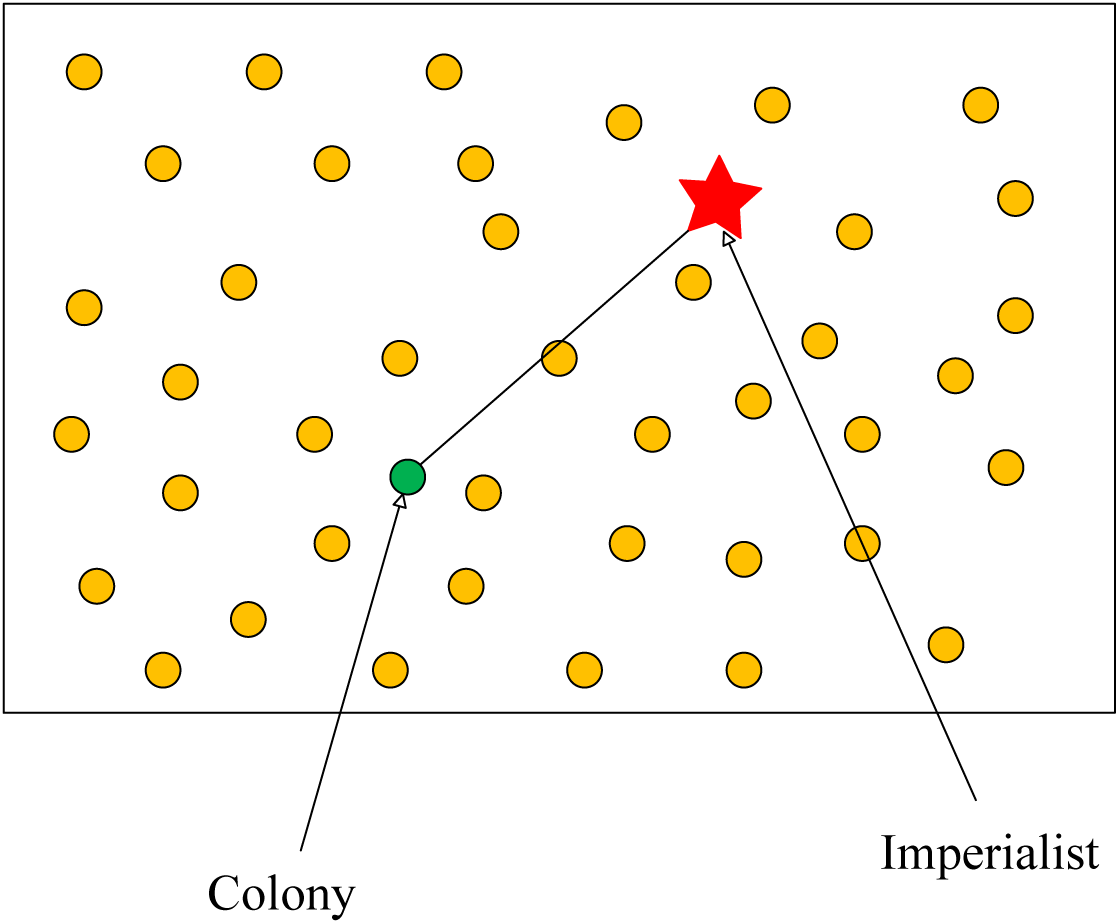
The initial empires generation [78]

The integration of this model with the neural network causes the error in the network to be defined as a cost function, and with the changes in weights and biases, the output of the network improves and the resulting error decreases [81]. The most important factors in training an ANN-ICA method are containing the number of countries, imperialists and decades and the number of neurons in the hidden layer which can be defined by different ways. One of the common ways for defining them is trial and error.

In the present study, inputs and outputs of each scenario where imported to the model. The number of countries, imperialists and decades have been defined by trial and error. For developing MLP model three architectures including 4-10-1, 4-14-1 and 4-18-1 (with 10, 14 and 18 neurons in the hidden layer, respectively).

#### 2.2.2 ANFIS

An adaptive neuro-fuzzy inference system (ANFIS) is a type of artificial neural network based on the Takagi-Sugeno fuzzy system [82,83]. This method was developed in the early 1990s. Since this system integrates neural networks and concepts of fuzzy logic, it can take advantage of the capabilities of both methods. It has nonlinear functions [82,83]. Figure 4 presents the architecture of the developed ANFIS model.

**Figure 4.**
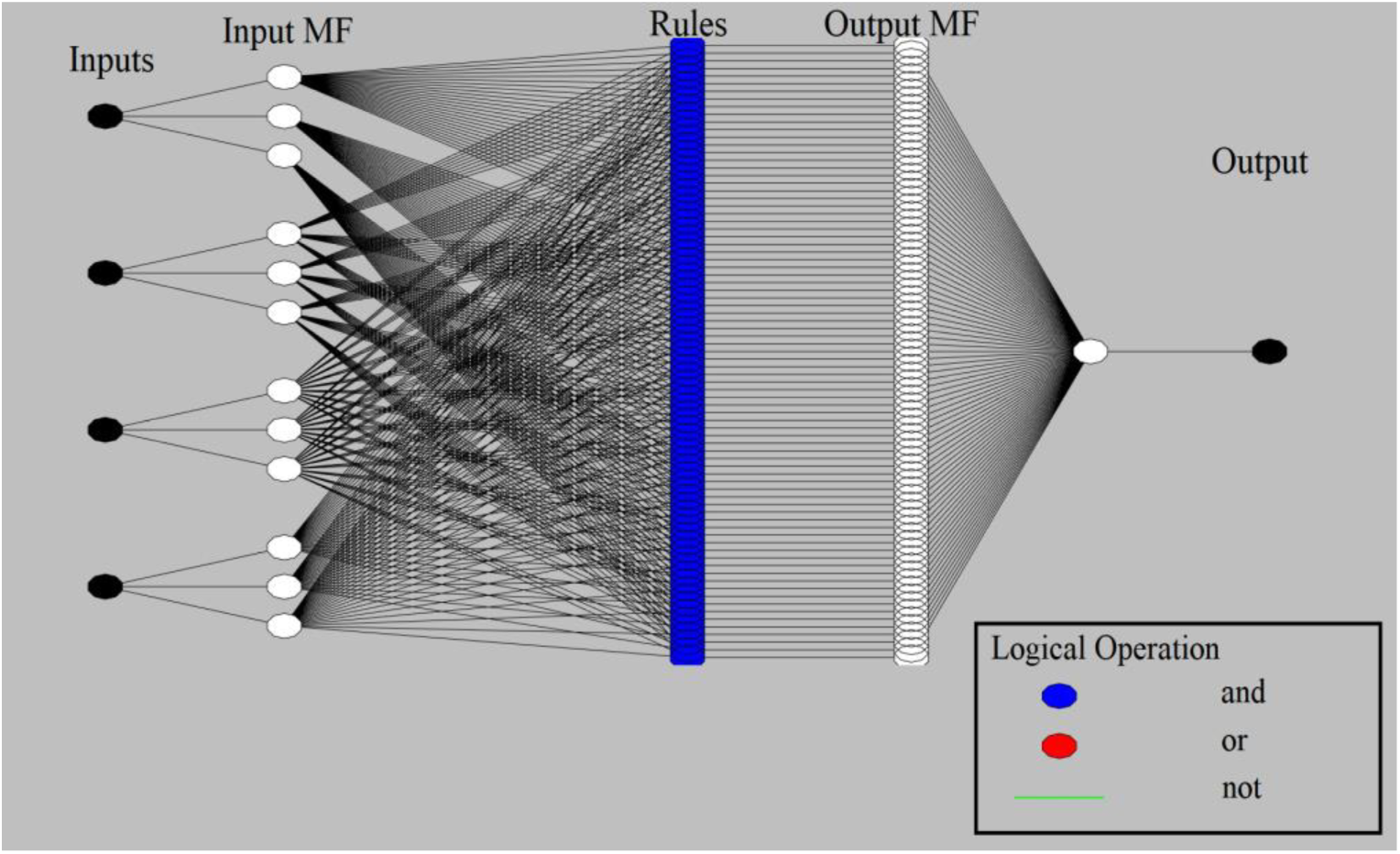
the architecture of the developed ANFIS

As is clear from Figure 4, ANFIS has five main layers [84–86]. The first layer is the inputs layer which takes the parameters and imports them to the model. This layer is also called as the input layer of the fuzzy system. The outputs of the first layer imports to the second layer and carries prior values of membership functions (MFs). Fuzzy rules are concluded from the nodes on second layer related degree of activity. The third layer normalizes the degree of activity of any rules. The fourth layer adopts the nodes and function and produces the outputs [87] and send them to the output layer. The important factors for determining the accuracy of ANFIS is the number and type of MFs, the optimum method and the output MF type [88–92].

ANFIS model was developed by ANFIS toolbox on MATLAB. Input parameters were independent variables of each scenario and the output variable was the number of cases or mortality rate. The ANFIS model was trained with three triangular, trapoizidal and gaussian MFs. This step was performed in order to select the best MF. The output membership function type selected linear type Because of its ability to further reduce of errors. Training of FIS was done with backpropagation optimum method and 0 value of error tolerance.

#### 2.2.3 Evaluation criteria

Evaluations were conducted by determination coefficient, root mean square error and mean absolute percentage error values. These factors compare the target and output values and calculates a score as an index for the performance and accuracy of the developed methods [93,94]. Table 2 presents the evaluation criteria equations.

**Table 2.**
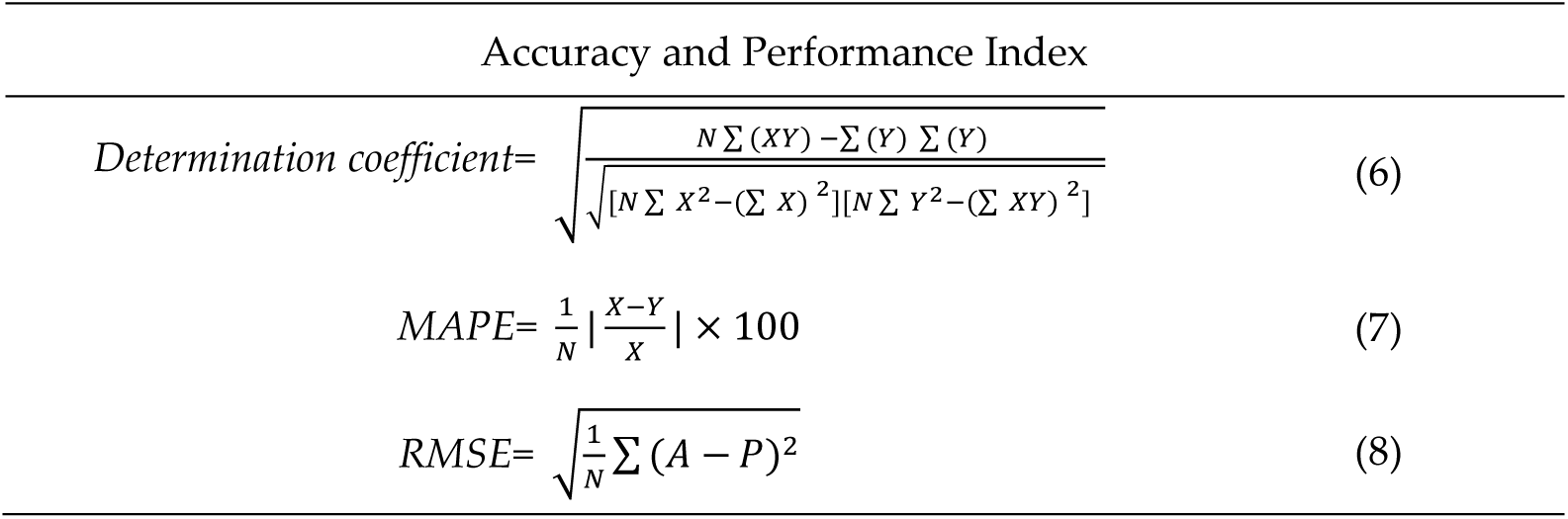
Model Evaluation metrics

where *N* is the number of data, *X* and *Y* are, respectively, predicted (output) and desired (target) values.

## 3. Results

The performance of the proposed algorithm is evaluated using both training and validation data. The training data are used to train the algorithm and define the best set of parameters to be used in ANFIS and MLP-ICA. After that, the best setup for each algorithm is used to predict outbreaks on the validation samples. Worth mentioning that due to the lack of adequate sample data to avoid the overfitting, the training is used to evaluate the model with higher performance.

### 3.1 Training results

The training step for ANFIS was performed by employing three MF types as described in tables 3 and 4. Table 3 presents the training results of ANFIS for COVID-19 cases and Table 4 presents the training results of ANFIS for COVID-19 mortality rate in Hungary. As is clear, results have been compared using evaluation metrics of RMSE and MAPE. According to Table 3, Gaussian MF type with MF number 3, BP optimum method and linear output MF provided the highest performance by the lowest value of RMSE. On the other hand, as is clear scenario 1 provided the lowest RMSE in comparison with scenario 2 for the selected MF. Therefore, it can be concluded that, scenario 1 is suitable for modeling COVID-19 cases in comparison with scenario 2.

**Table 3.** ANFIS training results (cases)

|  | MF type | No. of MFs | Optimum method | Output MF | Epoch No. | RMSE | MAPE |
| --- | --- | --- | --- | --- | --- | --- | --- |
| Scenario 1 | Triangular | 3 | Back-propagation | Linear | 233 | 248.06 | 38.44 |
|  | Trapezoidal | 3 | Back-propagation | Linear | 253 | 158.59 | 35.67 |
|  | **Gaussian** | **3** | **Back-propagation** | **Linear** | **388** | **119.42** | **39.95** |
| Scenario 2 | Triangular | 3 | Back-propagation | Linear | 266 | 250.26 | 37.75 |
|  | Trapezoidal | 3 | Back-propagation | Linear | 206 | 211.98 | 33.39 |
|  | **Gaussian** | **3** | **Back-propagation** | **Linear** | **285** | **182.76** | **34.6** |

According to Table 4, it can be claimed that Gaussian MF provided the lowest error and highest accuracy compared with other MF types for the prediction of mortality rate. Also it can be claimed that, for the selected MF type, scenario 2 provides the highest performance compared with scenario 1 for the prediction of mortality rate.

**Table 4.**
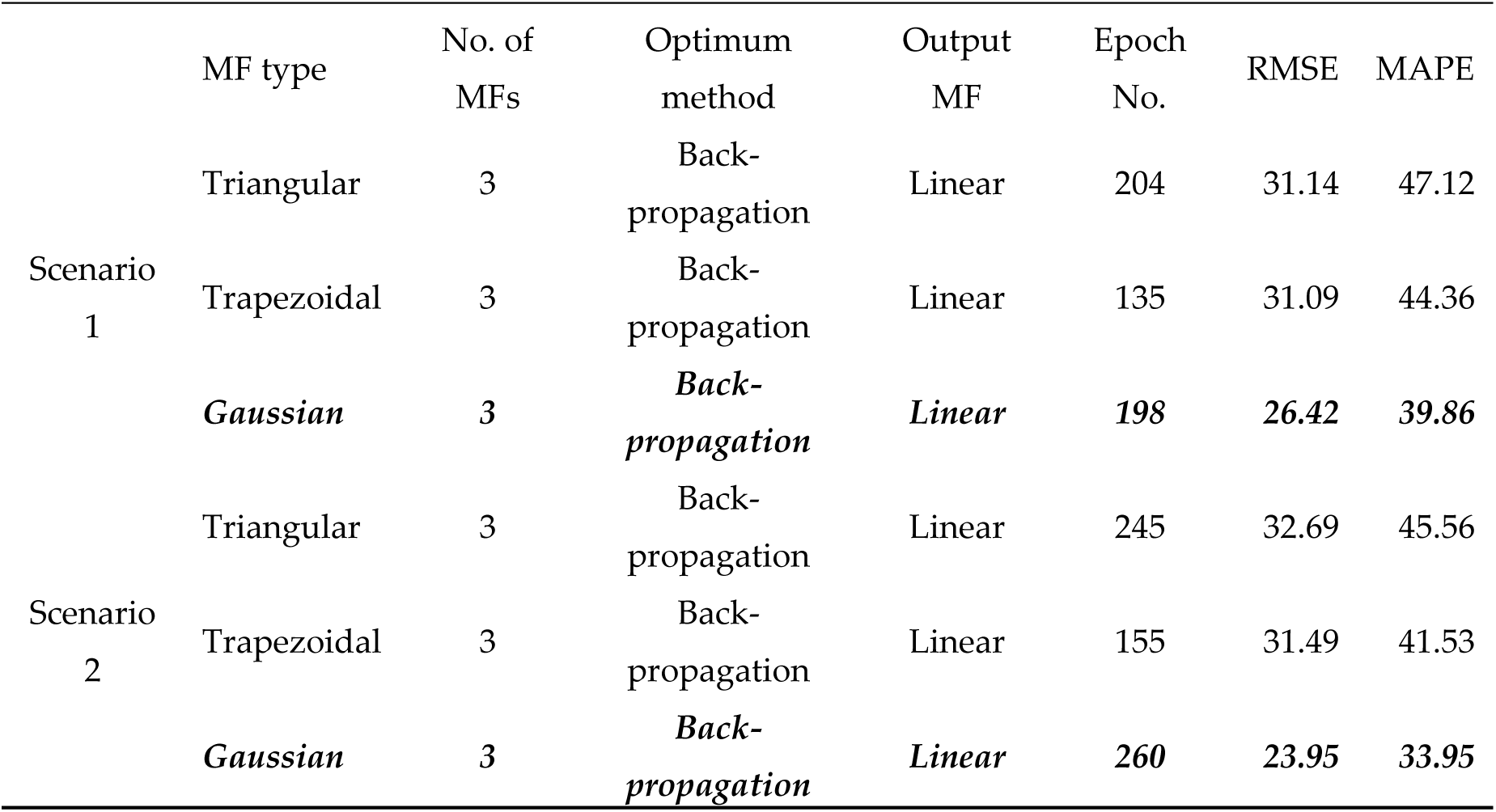
ANFIS training results (mortality rate)

Tables 5 and 6 presents results for the prediction of COVID-19 cases and mortality rate, respectively by MLP-ICA. According to Table 5, MLP architecture with 10 neurons in the hidden layer provided the highest accuracy for the prediction of COVID-19 cases in the presence of both scenarios. But, Scenario 2 provided higher accuracy than scenario 1 according to the lowest RMSE value.

**Table 5.** MLP-ICA training results (cases)

| Table 5. MLP-ICA training results (cases) |  |  |  |  |  |  |
| --- | --- | --- | --- | --- | --- | --- |
|  | No. of countries | No. of decades | No. of initial imperialists | No. of neurons | RMSE | MAPE (%) |
| Scenario 1 | *300* | *40* | *50* | *10* | *37.30* | *23.15* |
|  | 300 | 40 | 50 | 14 | 37.39 | 23.82 |
|  | 300 | 40 | 50 | 18 | 37.48 | 23.57 |
| Scenario 2 | *300* | *40* | *50* | *10* | *37.24* | *23.47* |
|  | 300 | 40 | 50 | 14 | 37.43 | 23.63 |
|  | 300 | 40 | 50 | 18 | 38.29 | 22.15 |

According to Table 6, neuron number 14 for scenario 1 and neuron number 18 for scenario 2 provided the highest accuracy in integrating by ICA method in comparison with other architectures. By comparing the evaluation criteria values, it can be concluded that, scenario 1 is more suitable than scenario 2 for the prediction of mortality rate using MLP-ICA.

**Table 6.**
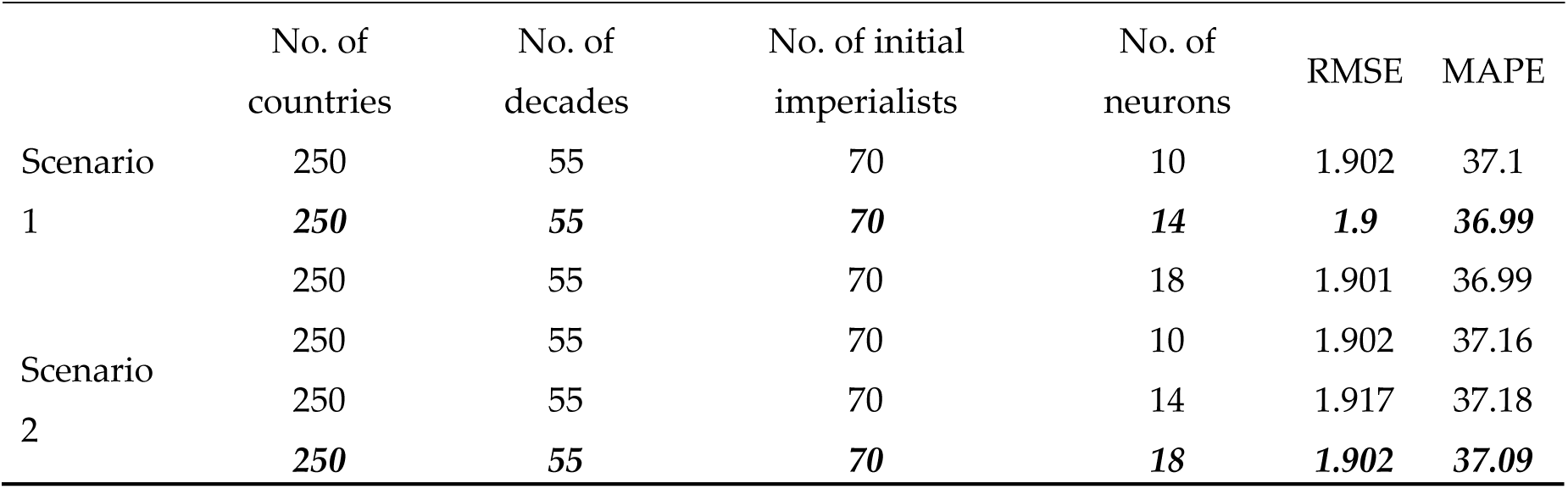
MLP-ICA training results (mortality rate)

Figure 3 presents the plot diagram for the selected models according to Table 3 to 6. In these figures, the vertical axis is “Target values” or in another word the actual values and the horizontal axis is the “predicted values” or in another word the output value of the model. In these plots, the dash-line is the 1:1 line. The distance of each point from the dash-line discusses the accuracy of the prediction. Such that, points on dash-line have the minimum error and increases the determination coefficient and points those that have distance from dash line increases the error and reduces the accuracy in accordance with its distance and reduces the determination coefficient.

As is clear from Figure 5, ANN provides a higher determination coefficient (0.9963, 0.9963, 0.9987 and 0.9987, respectively for the prediction of COVID-19 cases and COVID-19 mortality rate in the presence of scenario 1 and scenario (see the Figure 5)) than ANFIS. Also, the ability of ANN in the prediction of COVID-19 mortality rate is higher than that for the prediction of COVID-19 cases. But, ANFIS provides a little difference behavior than ANN. Such that, in ANFIS scenario 1 provides the highest determination coefficient for the prediction of COVID-19 cases but scenario 2 provides the highest correlation coefficient for the prediction of mortality rate (0.9689, 0.934, 0.8909 and 0.9427, respectively for the prediction of COVID-19 cases and COVID-19 mortality rate in the presence of scenario 1 and scenario (see the Figure 5)).

**Figure 5.**
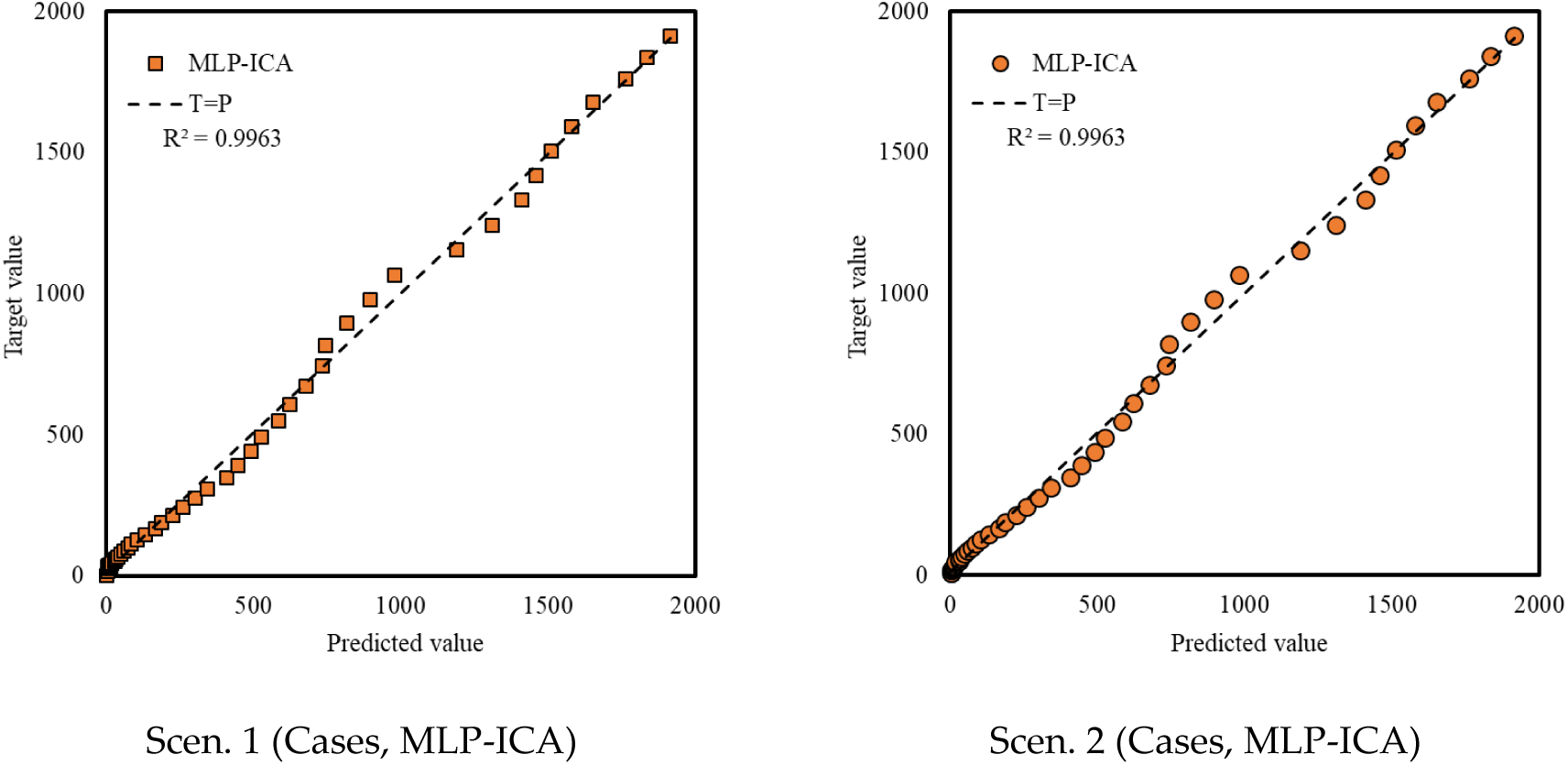
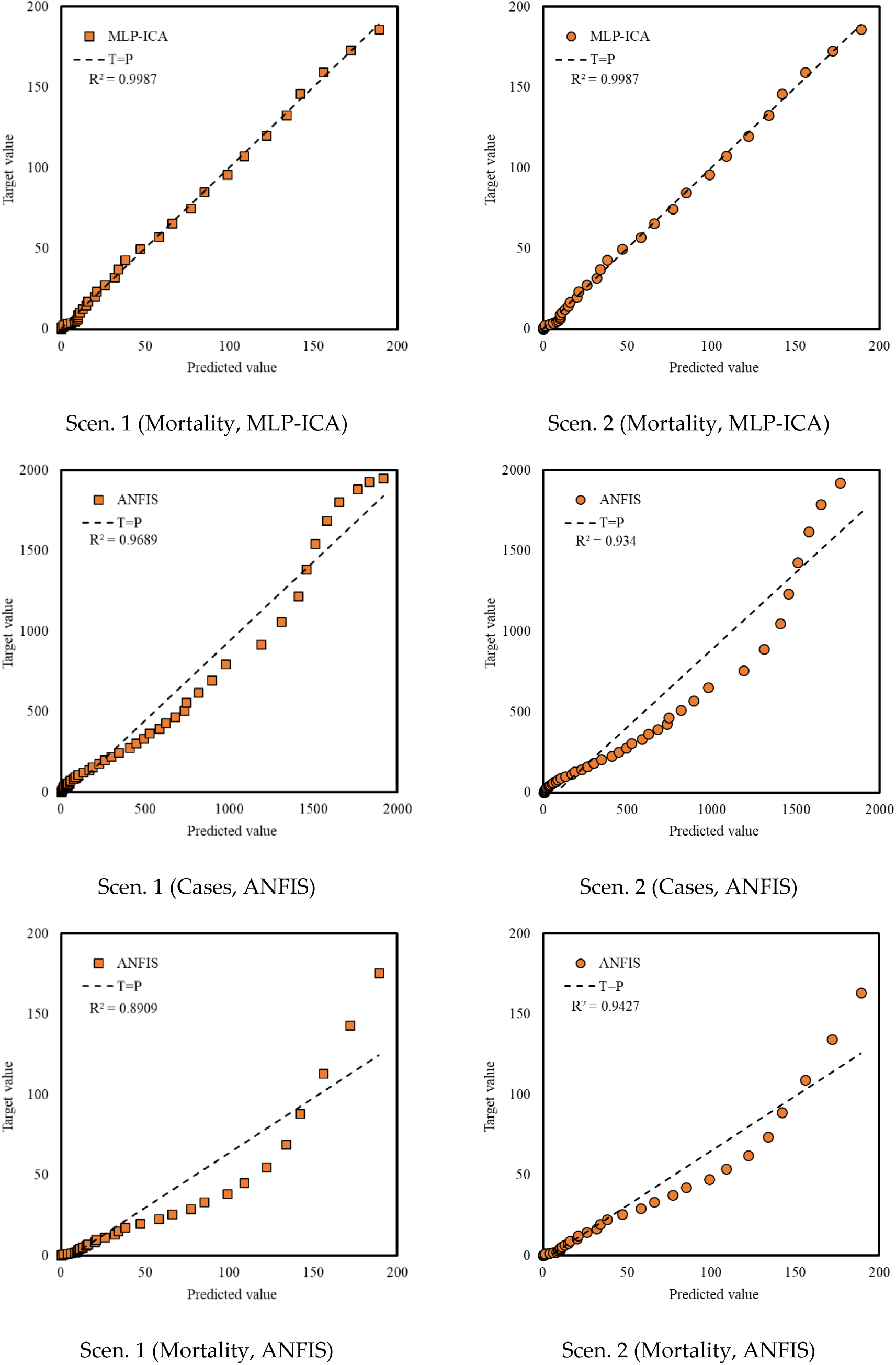
Plot diagram for the prediction of COVID-19 cases and mortality rate

Figure 6 presents the deviation from target values for the COVID-19 cases from 4-March to 19-April. According to Figure 6, MLP-ICA in the presence of scenario 2 provides lower deviation from target value followed by MLP-ICA in the presence of scenario1 than the ANFIS in the presence of both scenarios.

**Figure 6.**
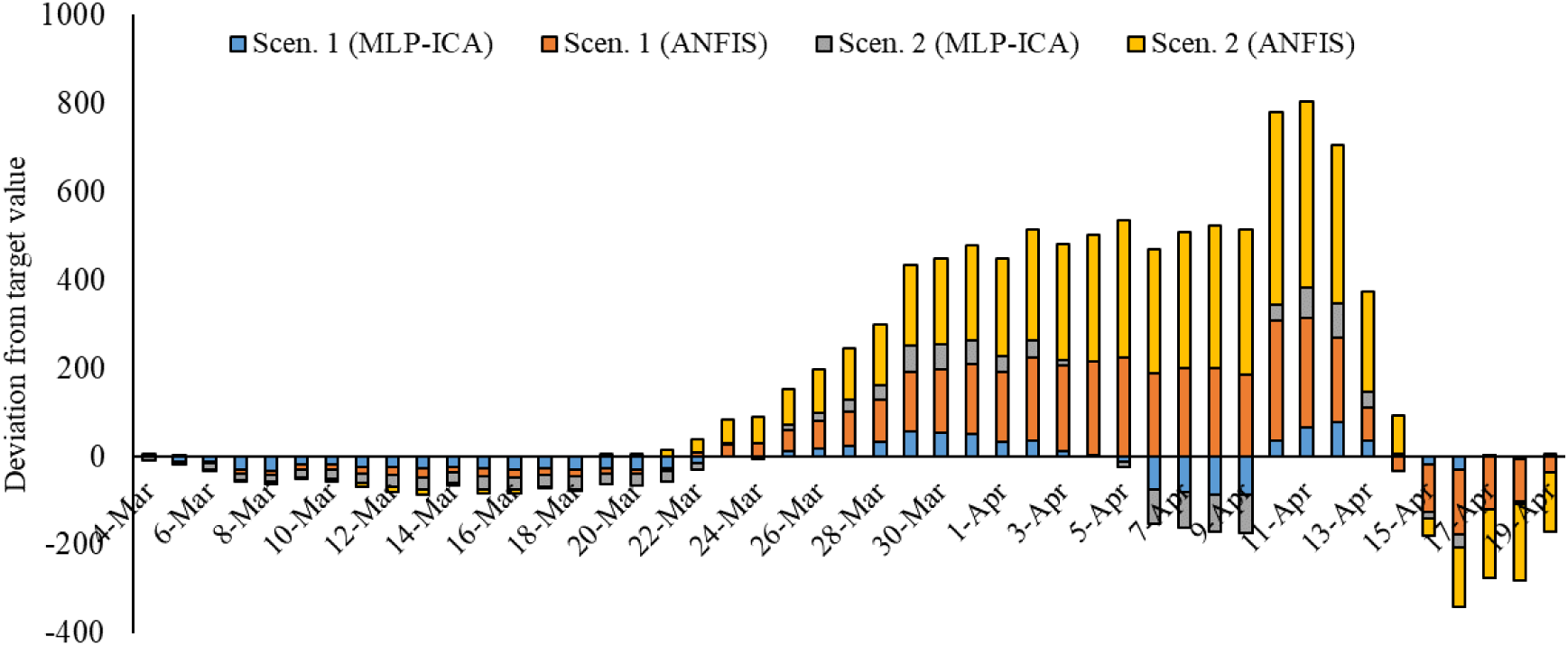
Deviation from target value (Cases)

Figure 7 presents the deviation from target values for the COVID-19 mortality rate from 4-March to 19-April. According to figure 7, MLP-ICA in the presence of scenario 1 provides lower deviation from target value followed by MLP-ICA in the presence of scenario2 than the ANFIS in the presence of both scenarios.

**Figure 7.**
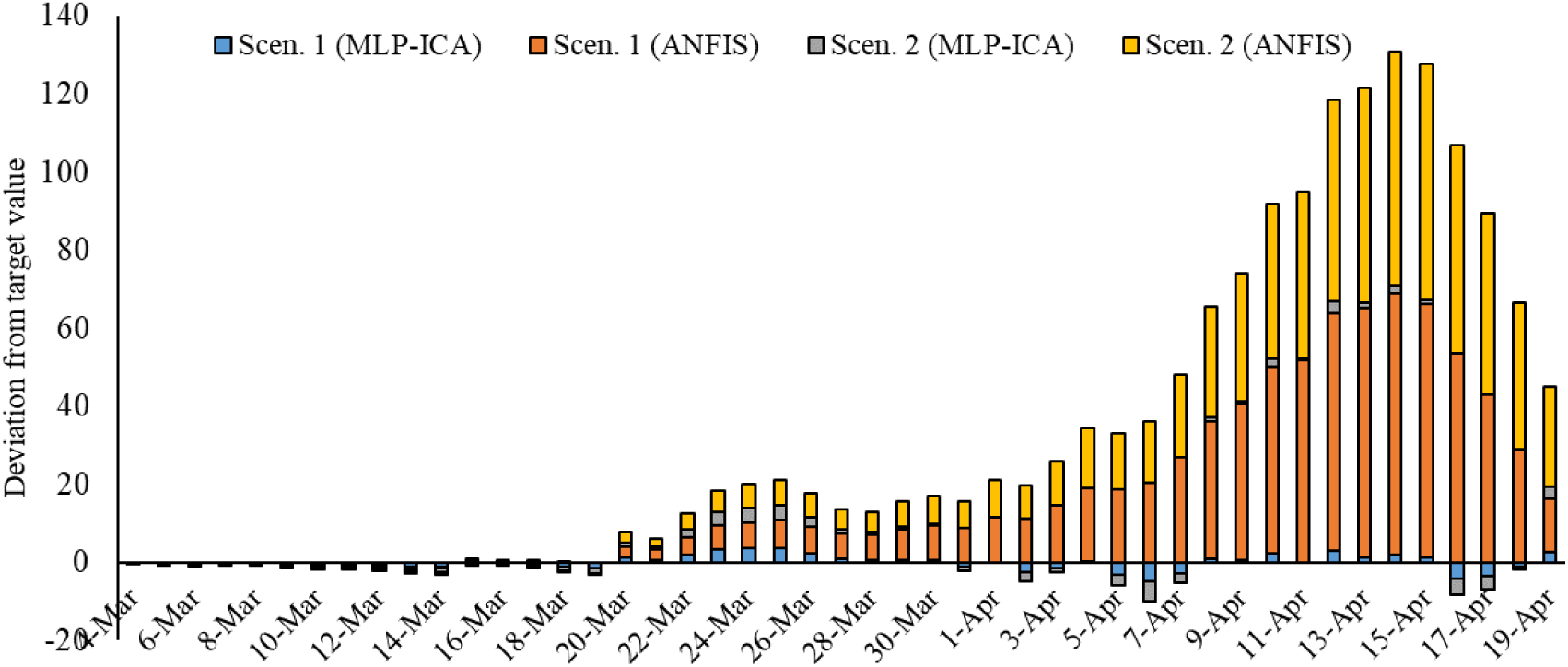
Deviation from target value (mortality rate)

By evaluating the modeling methods in the last sections, it was decided to select the MLP-ICA in the presence of scenario 2 for the prediction of COVID-19 outbreak and MLP-ICA in the presence of scenario 1 for the prediction of COVID-19 mortality rate in Hungary. Predictions were performed in two stages. First stage for total prediction and the second one for daily prediction. Figures 8 and 9 present total cases and total mortality rate, respectively, and Figure 10 and 11 present the daily Prediction of the results from 20-April to 30-July. Each figure has two sections including the reported statistic (from 24-March to 19-April) and the predicted by the selected model (from 20-April to 30-July).

**Figure 8.**
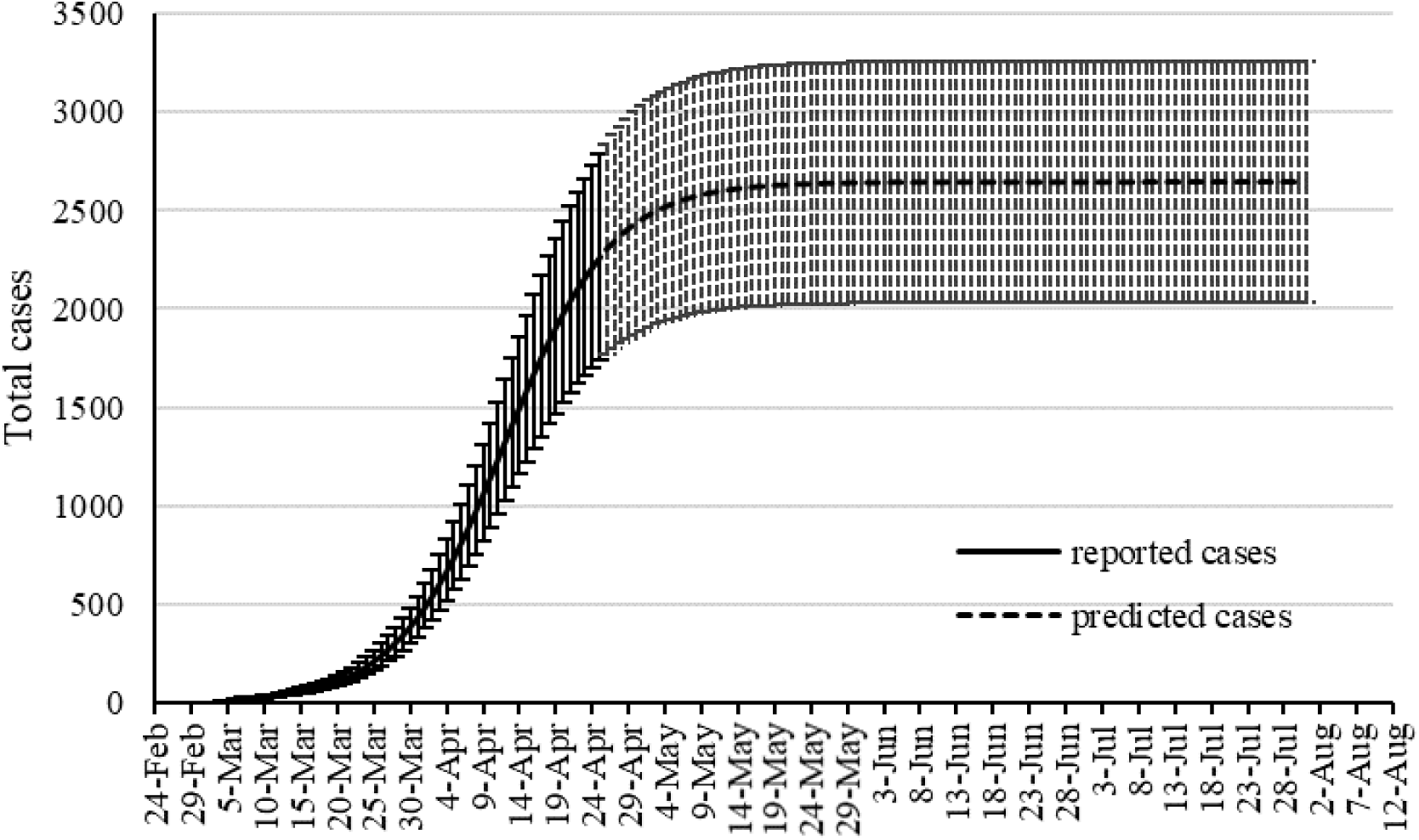
Total outbreak prediction for MLP-ICA

**Figure 9.**
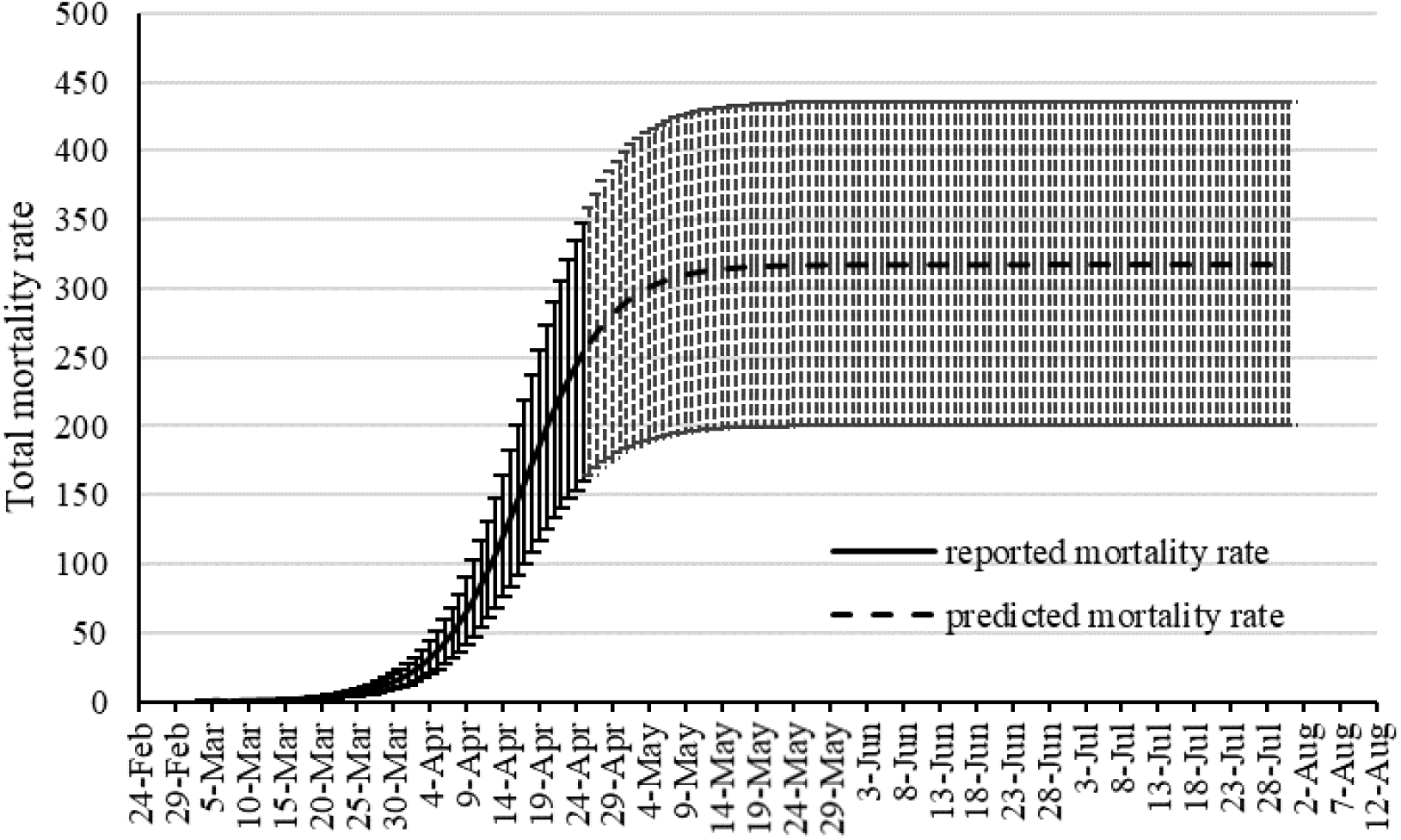
Total mortality rate prediction for MLP-ICA

**Figure 10.**
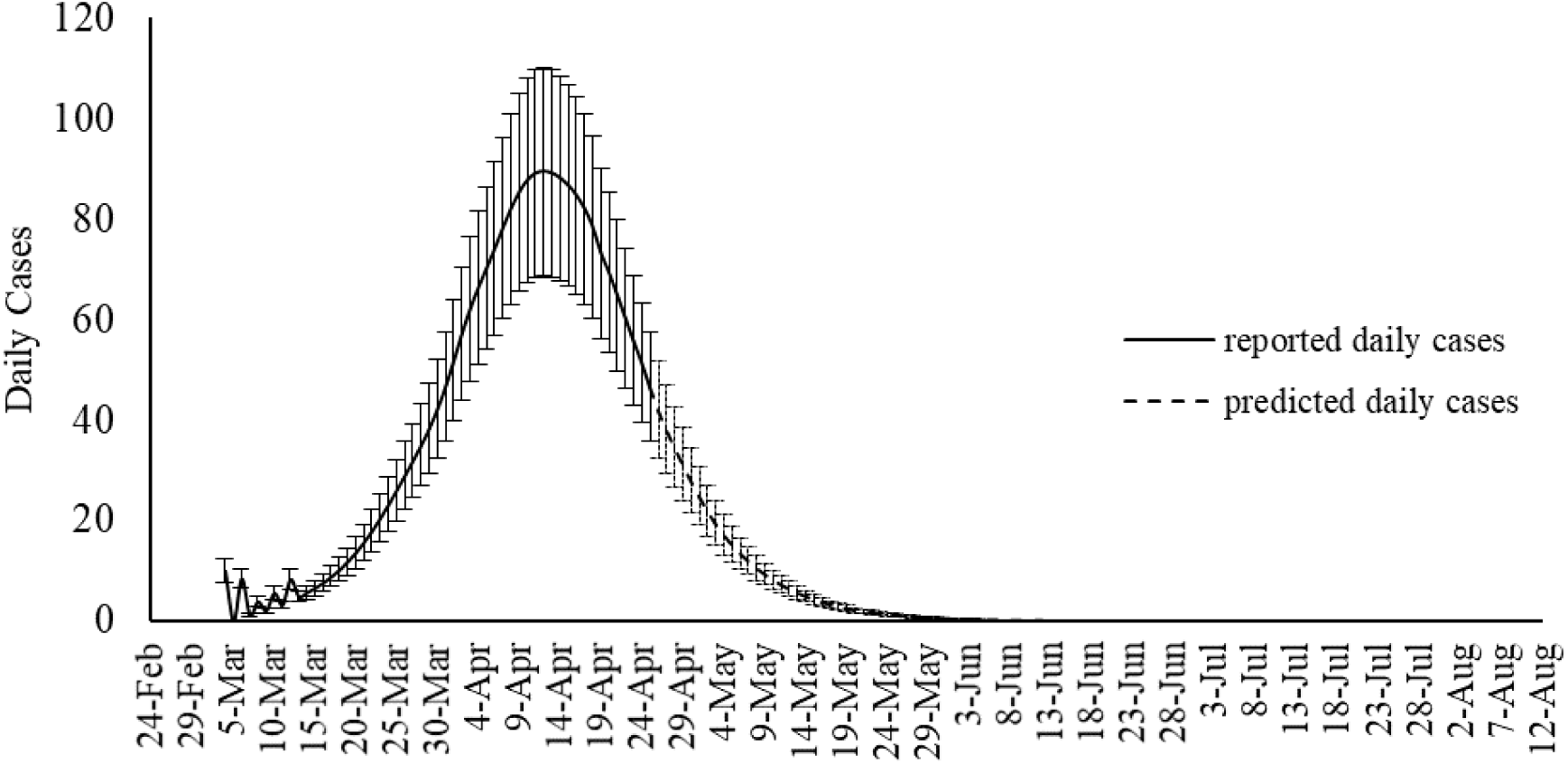
Daily outbreak prediction for MLP-ICA

**Figure 11.**
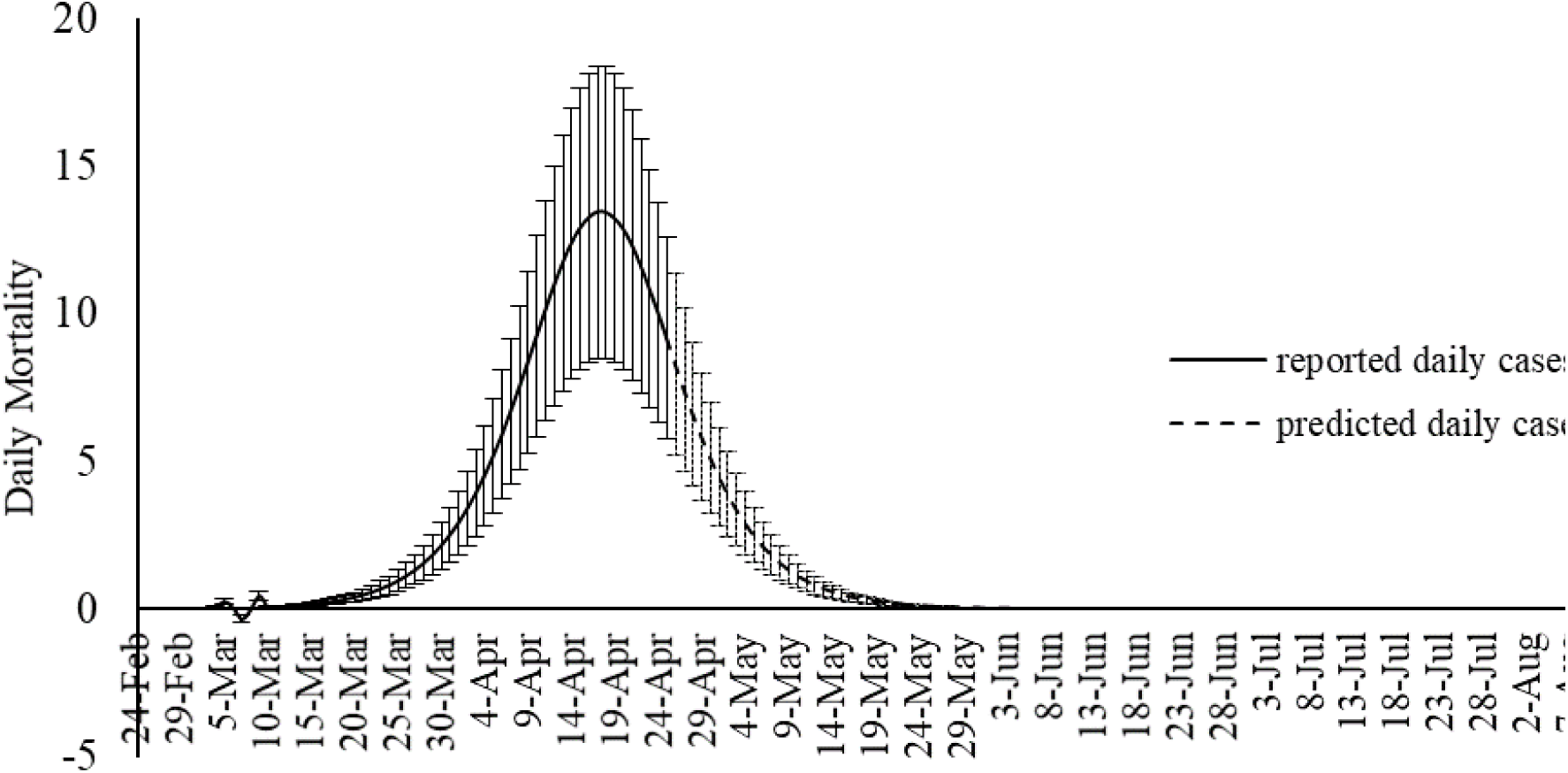
Daily mortality rate prediction for MLP-ICA

### 3.2 Validation

Table 7. represents the validation of the MPL-ICA and ANFIS models for the period of 20-28 April. The proposed model of MPL-ICA presented promising values for RMSE and determination coefficient for prediction of both outbreak and total mortality.

**Table 7.** Validation of the models for 9 days

|  | Total Cases |  | Total mortality rate |  |
| --- | --- | --- | --- | --- |
| For 20-28 April | MPL-ICA | ANFIS | MLP-ICA | ANFIS |
| RMSE | 167.88 | 194.10 | 8.32 | 15.25 |
| Determination coefficient | 0.9971 | 0.9563 | 0.9986 | 0.7491 |

## 5. Discussions

This approach outperforms commonly used prediction tools in the case of Hungary. More work is required if this technique is adequate in all cases and for different population types and sizes. Nonetheless, the learning approach could overcome imperfect input data. Incomplete catalogs can occur because infected persons are asymptotic, not tested, or not listed in databases. Tests in a closed environment such as large aircraft carriers in France and the US have shown that up to 60% of the infected personnel were asymptomatic. Of course, military personals are not representative of large and mixed populations. Nonetheless, it shows that false negatives can be abundant. In emerging countries, access to laboratory equipment required for testing is extremely limited. This will introduce a bias in the counting. Finally, it is unclear if all the cases are registered. In the UK for example, it took public pressure for the government to make the casualties in retirement hospices known. And there is still doubt in the community that China produced complete data for political reasons. At the same time, national governments and local administrations implemented containment measures such as confinement and social distancing. These actions have a huge impact on transmissions and thus on cases and casualties. Access to modern medical facilities is also a parameter that mainly affects the number of casualties. All these aspects will affect traditional estimation procedures whereas learning algorithms might be able to adapt, especially if multiple datasets are available for a given region. Not only can our approach outperform the commonly used SIR but it requires fewer input data to estimate the trends. While we provide successful results for Hungarian data we need to further test these novel approaches on other databases. Nonetheless, the presented results are promising and should incite the community to implement these new tools rapidly.

## 4. Conclusions

Although SIR-based models been widely used for modeling the COVID-19 outbreak, they include some degree of uncertainties. Several advancements are emerging to improve the quality of SIR-based models suitable to COVID-19 outbreak. As an alternative to the SIR-based models, this study proposed machine learning as a new trend in advancing outbreak models. The machine learning approach makes no assumption on the pandemic and spread of the infection. Instead it predicts the time series of the infected cases as well as total mortality cases.

In this study the hybrid machine learning model of MLP-ICA and ANFIS are used to predict the COVID-19 outbreak in Hungary. The models predict that by late May the outbreak and the total morality will drop substantially. Based on the results reported here, and due to the complex nature of the COVID-19 outbreak and variation in its behavior from nation-to-nation, this study suggests machine learning as an effective tool to model the outbreak. Two scenarios were proposed. Scenario 1 considered sampling the odd days and Scenario 2 used even days for training the data. Training the two machine learning models ANFIS and MLP-ICA were considered for the two scenarios. A detailed investigation was also carried out to explore the most suitable number of neurons. Furthermore, the performance of the proposed algorithm is evaluated using both training and validation data. The training data are used to train the algorithm and define the best set of parameters to be used in ANFIS and MLP-ICA. After that, the best setup for each algorithm is used to predict outbreaks on the validation samples. The validation is performed for nine days with promising results which confirms the model accuracy. In this study due to the lack of adequate sample data to avoid the overfitting, the training is used to choose and evaluate the model with higher performance. In the future research, as the COVID-19 progress in time and with the availability of more sample data further testing and validation can be used to better evaluate the models.

Both models showed promising results in terms of predicting the time series without the assumptions that epidemiological models require. Both machine learning models, as an alternative to epidemiological models, showed potential in predicting COVID-19 outbreak as well as estimating total mortality. Yet, MLP-ICA outperformed ANFIS with delivering accurate results on validation samples. Considering the availability of a small amount of training data, further investigation would be essential to explore the true capability of the proposed hybrid model. It is expected that the model maintains its accuracy as long as no major interruption occurs. For instance, if other outbreaks would initiate in the other cities, or the prevention regime changes, naturally the model will not maintain its accuracy. For the future studies advancing the deep learning and deep reinforcement learning models is strongly encouraged for comparative studies on various ML models for individual countries.

Nomenclatures

|  |  |  |  |
| --- | --- | --- | --- |
| Multi-layered perceptron | MLP | Susceptible-infectious-susceptible | SIS |
| Adaptive network-based fuzzy inference system | ANFIS | Mean square error | MSE |
| Call data record | CDR | Artificial intelligence | AI |
| Classification and regression tree | CART | Artificial neural network | ANN |
| Evolutionary algorithms | EA | Susceptible-exposed-infected-recovered | SEIR |
| Imperialist competitive algorithm | ICA | Random Forest | RF |
| Maternally-derived-immunity-susceptible-exposed-infected-recovered-susceptible | MSEIRS | Maternally-derived-immunity-susceptible-exposed-infected-recovered | MSEIR |
| Maternally-derived-immunity-susceptible-infected-recovered | MSIR | Susceptible-exposed-infected-susceptible | SEIS |
| Membership function | MF | Machine learning | ML |
| Particle swarm optimization | PSO | Neural Networks | NN |
| Susceptible-infected-recovered | SIR | Root mean square error | RMSE |
| Susceptible-infected-recovered-deceased | SIRD | Gaussian | Gauss. |

## Data Availability

Data is online from WHO

## Acknowledgments

We acknowledge the financial support of this work by the Hungarian State and the European Union under the EFOP-3.6.1-16-2016-00010 project and the 2017-1.3.1-VKE-2017-00025 project.

## Author contribution

Conceptualization, Amir Mosavi; Data curation, Gergo Pinter and Imre Felde; Formal analysis, Gergo Pinter and Imre Felde; Investigation, Sina F. Ardabili; Methodology, Sina F. Ardabili, Amir Mosavi and Pedram Ghamisi; Resources, Imre Felde, Amir Mosavi and Pedram Ghamisi; Supervision, Imre Felde, Amir Mosavi and Pedram Ghamisi; Validation, Sina F. Ardabili and Amir Mosavi; Visualization, Sina F. Ardabili; Writing - original draft, Amir Mosavi; Writing - review & editing, Amir Mosavi and Pedram Ghamisi.

